# Healthcare worker mask reuse in a global pandemic: Using idle resources to create an inexpensive, scalable, and accessible UV system for N95 sterilization

**DOI:** 10.1101/2020.04.19.20070870

**Authors:** Rachel M. Gilbert, Michael J. Donzanti, Daniel J. Minahan, Jasmine Shirazi, Christine L. Hatem, Brielle Hayward-Piatkovskyi, Allyson M. Dang, Katherine M. Nelson, Kimberly L. Bothi, Jason P. Gleghorn

**Affiliations:** Departments of Biomedical Engineering, University of Delaware, Newark, DE USA 19716; Departments of Biological Sciences, University of Delaware, Newark, DE USA 19716; Departments of Chemistry and Biochemistry, University of Delaware, Newark, DE USA 19716; Departments of Chemical and Biomolecular Engineering, University of Delaware, Newark, DE USA 19716; Departments of UD Global Engineering, University of Delaware, Newark, DE USA 19716

## Abstract

As the current COVID-19 pandemic illustrates, not all hospitals and other facilities are equipped with enough personal protective equipment to meet the demand in a crisis. Healthcare workers around the world utilize N95 masks to protect themselves and their patients, yet during this global pandemic they are forced to re-wear what is intended to be single-use masks. This poses significant risk to these healthcare workers along with the populations they are trying to protect. Ultraviolet germicidal irradiation (UVGI) has been validated previously as a way to effectively sterilize these masks between use, however, not all facilities have access to the high cost commercial UV-C lamp sterilization equipment. However, UV-C bulbs are sitting idle in biosafety cabinets (BSCs) at universities and research facilities around the globe that have been shuttered to slow the spread of COVID-19. These bulbs may also be available in existing medical centers where infectious diseases are commonly treated. Therefore, we have developed a method to modify existing light fixtures, or create custom light fixtures compatible with new or existing common UV-C bulbs. **This system is scalable and can be created for less than 50 US dollars, on site, at the point of need, and leverages resources that are currently untapped and sitting unused in public and private research facilities**. The freely-accessible design can be easily modified for use around the world. Hospitals can obtain this potentially life-saving UVGI resource with minimal funds, via collaboration between research facilities to obtain the UV-C meters and limited availability UVGI bulbs. While mask reuse is not ideal, we must do what we can in emergency situations to protect our frontline healthcare workers and the communities they serve.

## INTRODUCTION

Front line health care workers (HCW) are critical to the care and treatment of individuals with the novel coronavirus, SARS-CoV-2, or disease known as COVID-19. In addition to the needed beds and ventilators, personal protective equipment (PPE), particularly filtering facepiece respirators (FFRs), are essential to ensure the health and safety of not only trained doctors, nurses, and emergency response personnel, but also other hospital staff who play an important role in cleaning, disinfecting, and preparing spaces for patient care. Additionally, whereas much of the focus is on large hospitals and current hotspots without access to FFRs, also called N95 masks, SARS-CoV-2 will spread and affect residential facilities and rural clinics around the globe. These communities face additional challenges with limited resources and larger logistical obstacles to obtaining FFRs. And at the time of publication, shipping worn FFRs to localized centers for hydrogen peroxide vapor (HPV, also written as H_2_O_2_ vapor) sterilization was only available in very limited locations in the United States of America (U.S.) [1–3]. HPV sterilization is a U.S. Food & Drug Administration-approved method for N95 mask sterilization, and manufacturing and deployment of these systems is currently underway; however, the operational and coordination challenges associated with even localized deployment of HPV centers for N95 sterilization are significant. This is evidenced by contemporary reports of front-line HCWs being issued N95 masks for continuous (re)use over week-long time periods [4–6]. Additionally, Nebraska Medicine has initiated a U.S. Center for Disease Control (CDC) approved on-site ultraviolet germicidal irradiation (UVGI) sterilization system for N95 mask decontamination [7]. UVGI has been demonstrated to be effective at quickly sterilizing FFRs for viruses like the novel SARS-CoV-2, and for multiple cycles of sterilization [8–10]. Whereas UVGI sterilization has important limitations as discussed herein, these methods currently <u>are</u> being deployed as emergency procedures during the SARS-CoV-2 pandemic. The Nebraska Medicine protocol uses an operating room UVGI sterilization system to set-up N95 mask decontamination, a system that many smaller clinics, rural hospitals and residential facilities may not have. We document procedures to build a similar type of UVGI irradiation platform with off-the-shelf components from the hardware store and UVGI bulbs sold online or from biosafety cabinets (class I, II, or III) that are ubiquitously found throughout academic research and industrial centers around the world. **This system is scalable and can be created for less than 50 US dollars, on site, at the point of need, and leverages resources that are currently untapped and sitting unused in public and private research facilities that have shut down during the SARS-CoV-2 pandemic**. Hospitals can obtain this potentially life-saving UVGI resource with minimal funds, via collaboration between research facilities to obtain the UV-C meters and limited availability UVGI bulbs.

### Demand and need for FFRs/N95 masks during 2019-2020 coronavirus pandemic

The SARS-CoV-2 pandemic is expected to continue to increase the burden on healthcare providers. As the number of cases increase, hospitals will continue to be stretched to their limits in terms of supplies and labor. There are 6,146 hospitals in the United States, 5,198 of which are classed as community hospitals (the remainder are federal government hospitals (209), nonfederal psychiatric hospitals (616), and other hospitals (123)) [11]. Community hospitals are those hospitals that can be accessed by the general public and include short-term general and specialty hospitals. Of the community hospitals, there are two major classifications: rural and urban. There are 3,377 urban community hospitals that serve approximately 106,000 square miles (∼84% of the population) while there are 1,821 rural community hospitals that serve approximately 3.4 million square miles (∼16% of the U.S. population) [12]. These numbers do not include urgent care centers, doctor’s offices, and other non-hospital medical sites on the front line of a pandemic response.

In contrast, many countries across the Global South rely heavily on a limited number of fully-resourced hospitals in urban centers, with varying degrees of professional healthcare access in rural areas. Current data on health facilities is difficult to find in many developing countries, however, we can find examples of the resource constraints. Kenya, for example, has 842 public and private hospitals serving over 53 million people [13,14]. Just 24 of these facilities are classified as county and national referral hospitals, and large teaching and private hospitals. The remainder of Kenya’s over 11,000 health facilities include smaller clinics, dispensaries, health centers, maternity wards and nursing homes, not to mention thousands of volunteer health care workers in rural communities. There are currently dramatic shortfalls in protective equipment in countries with relatively robust healthcare services, like the U.S., and these resources are even more precious in developing communities where fewer doctors and nurses are serving larger populations. According to the World Health Organization (WHO), “Africa suffers more than 22% of the global burden of disease but has access to only 3% of healthcare workers, and less than 1% of the world’s financial resources [15].” Losing a single doctor during this pandemic can have a detrimental impact on already strained healthcare systems across the continent [16]. Even in the U.S., rural clinics and hospitals serve patient populations sometimes across hundreds of miles, and in some areas, there is a single doctor for several thousand square miles [17]. Ensuring frontline HCWs around the world are protected as best possible is not only ethical, but imperative.

Challenges exist for both urban and rural healthcare facilities globally. Urban hospitals, while often having access to more resources due to the larger population they serve, experience a strain on their resources during a pandemic precisely because of the significantly larger numbers of people they need to urgently treat. Conversely, rural clinics and hospitals around the globe often have less funds to operate and face additional logistical challenges to provide patient access to care. Facilities cannot afford to have staff become ill and lead to a decrease in the number of HCW to treat patients, and therefore need proper FFRs to protect themselves. Additionally, HCWs can potentially spread infection if not properly equipped with essential FFRs, or if forced to reuse potentially contaminated masks. Many HCWs are currently facing the options of not wearing an essential mask, constructing makeshift FFRs with limited efficacy, or re-wearing a soiled mask - the majority are choosing the latter. This is an unprecedented world-wide shortage of life-saving equipment that our HCWs and beyond need in order to continue serving their communities. In addition to traditional healthcare workers, essential personnel working in pharmaceuticals, dentistry, custodial services, delivery services and law enforcement also require protection while they keep operations afloat. A global shortage of N95 masks is expected to persevere due to supply chain challenges, especially for one essential component: the melt-blown polypropylene fabric material that filters infectious diseases like the coronavirus during inhalation by the wearer [18]. Distributed systems for N95 sterilization are needed to keep up with demand.

The CDC estimated that a 42-day influenza outbreak in the U.S., which represents just 4.25% of the global population, could require over 90 million N95 masks for HCWs alone [19]. This would scale to almost 800 million FFRs in a year. A model of a hypothetical influenza pandemic predicted 1.7 to 7.3 billion respirators would be required if only 20-30% of the U.S. population were to be infected [20]. This does not account for non-HCWs, such as law enforcement officers and other essential personnel who may require respiratory protection. Given the uncertain nature of this pandemic and demonstrated logistical challenges in obtaining adequate resources, it is reasonable to assume that need will far exceed the value given in this projection and that demand will only grow.

### Sterilization and reuse of FFRs/N95 mask

N95 masks are designed and manufactured for single-use applications. The inability to scale N95 manufacturing at the rate needed to meet current demand during the SARS-CoV-2 pandemic has necessitated the reuse of N95 respirators among healthcare workers. Work has shown that pathogens such as viruses can contaminate and exist for extended periods of time on the outer surface of N95 masks [21]. Beyond the risk to HCWs in storing and reusing what is intended to be single-use PPE, other at-risk patients could be exposed to the virus when consulting with a HCW reusing their PPE that was previously used with a COVID19-positive patient. It should be noted that simply re-donning N95 masks alone poses serious risk to the user due to loss of strap elasticity, nose fit, and therefore, mask integrity, after repeated re-donning. Due to need, re-donning is already occurring, and these masks have the potential to be contaminated with viral particles, risking further spread of virus.

Sterilization of N95 masks must be considered carefully because improper sterilization can also give users a false sense of security in addition to compromising mask integrity. A variety of options have recently been developed to allow for sterilization between uses including HPV sterilization, UVGI treatment, and the applications of heat/humidity/washing [22]. The recently established N95DECON website (https://www.n95decon.org) gives a good summary of these methods, including current understandings and limitations to consider for each method. The FDA has approved hydrogen peroxide-based sterilization via one U.S. company, Battelle, in the state of Ohio. While an excellent resource, the procedure relies on shipment of contaminated masks to Ohio for sterilization [23,24] and/or the production of such HPV sterilization equipment for regional deployment. The ability for hospitals and care workers to sterilize their own masks in minutes, as opposed to days, is a great advantage in the use of UVGI.

Successful implementation of UVGI in a hospital setting is already being utilized by Nebraska Medicine [7]. Unfortunately, their system requires two surgical suite UVGI towers, with each costing in excess of US$20,000, which not all hospitals have available. Herein, we developed a UVGI lamp set-up that provides the capability for hospitals and local regional centers that do not have access to operating room UVGI towers to implement their own N95 mask sterilization system. These hospitals can make use of our proposed UVGI lamp, along with the work flow developed by Nebraska Medicine, to sterilize N95 masks in their own centers. Our UVGI lamp is accessible, inexpensive, requires little expertise to construct and operate, and repurposes existing UV-C bulbs not currently in use. The system takes advantage of common parts available at any hardware store. Once implemented, this method allows for high throughput, quick sterilization cycles which should allow for safer re-donning of N95 masks.

### UVGI as a method for FFR/N95 sterilization

UVGI systems have been used throughout the healthcare industry to sterilize work environments such as surgical suites, equipment, and ambulances. Single stranded RNA (ssRNA) viruses, like the novel SARS-CoV-2, are especially susceptible to UV sterilization [8]. Previous work has shown that UVGI systems can also be used to sterilize N95 masks by reducing the viability of the influenza virus, also a ssRNA virus, by 3 log [9]. A very recent small study has shown that UV is capable of sterilizing N95 mask fabric contaminated with SARS-CoV-2 [25]. There is variable effectiveness of UVGI depending on the mask manufacturer, the different materials of the mask (polypropylene filter *vs* rubber strap), and the medium in which the virus resides (in liquid, in air, on surface). UVGI sterilization also runs the risk of damaging the materials of the N95 masks which can compromise the integrity of the mask and its usefulness in filtering particles and acting as an effective piece of PPE. A variety of studies have looked at the effect of UVGI on mask integrity [26,27], even with repeated exposure [10,22,28–30], and have found no significant increase in viral penetration, nor decreases in mask stability, even at UVGI doses >10,000x the required dose to effectively reduce influenza infectivity [31]. In other studies, three UVGI cycles of 1.6-2.0 mW/cm^2^ for 15 min did not cause significant changes in respirator fit [28], and there was no change in filtration performance [29]. However, with UVGI sterilization of N95 masks, careful monitoring of UV dosage and the number of times a single mask is sterilized is important to minimize damaging the integrity of the mask. **Table 1** provides a summary of current literature related to UVGI sterilization of N95 mask, mainly in the context of influenza. Importantly, the CDC has approved the protocol from Nebraska for UVGI sterilization during the COVID-19 pandemic.

**Table 1:**
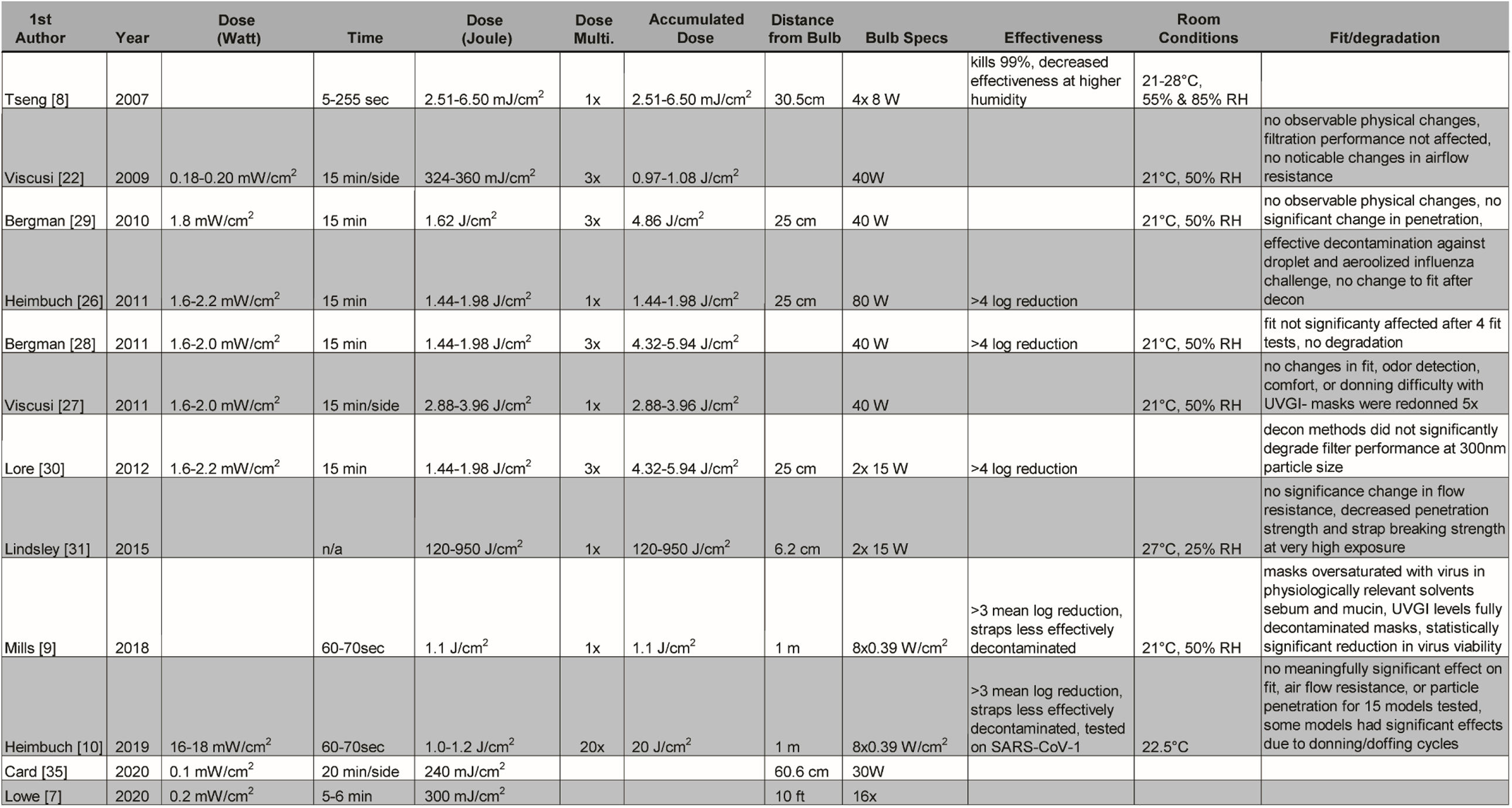
Summary of literature utilizing UVGI to sterilize FFRs/N95 masks

UVGI inactivates viruses by damaging their nucleic acids and, to a lesser extent, their protein capsid. UV-C wavelengths (100-280nm) have the highest sterilization efficiency because the maximum absorption wavelength is 260 nm and 280 nm for nucleic acids and proteins, respectively [32]. While little is known about the novel coronavirus SARS-CoV-2, comparisons between SARS-CoV-2 and SARS-CoV-1 can be helpful in estimating conditions in which SARS-CoV-2 may persist. As mentioned, ssRNA viruses, like SARS-CoV-2, are the most susceptible type of virus to UVGI, which is important for understanding the potential success of UVGI treatment [8]. SARS-CoV-2 can persist on plastic and stainless steel for up to 3 days, although at 73 hours virus titer had decreased 3-fold, and no viable SARS-CoV-2 was measured on cardboard after 24 hours [33]. How long this virus can remain viable on PPE has yet to be studied, and is likely a function of room humidity, contaminating fluids, and mask materials and construction.

### SOLUTION: Inexpensive, scalable, and accessible UVGI irradiation system for FFR/N95 sterilization for front-line medical workers

We propose a collaboration between public and private research institutions and hospitals to increase the access to UVGI sterilization. Currently, UVGI bulbs are in limited supply; however, most laboratory bio-safety cabinets (BSCs) are equipped with UVGI bulbs, and thousands of these bulbs are currently sitting idle, as research has been mainly halted to slow the spread of the virus. Besides research facilities, these bulbs might also be available in existing hospital settings like tuberculosis wards where they can be used for disinfection via passive fixtures or air handling units [34]. While others have proposed sterilizing FFRs within these BSCs directly [35], this requires transport of the masks to and from the research institutions, and staff to run the sterilization cycles. Our approach advances previous work and that from Nebraska Medicine and allows medical sites to create their own sterilization system, utilizing the idle bulbs, by retrofitting or creating custom light fixtures with off-the-shelf parts available at any retail hardware store.

We have developed step-by-step build plans to create UVGI light fixtures. These build plans can be downloaded from our website: https://www.gleghornlab.com/uvgi-sterilization. There are multiple options for fixture assembly, depending on availability of materials, which include:

1. Modification of premade commercial light fixtures to fit new or existing UVGI bulbs
  - Use an existing ceiling light fixture
  - Modify a hanging ceiling fixture
2. Create a custom fixture from off-the-shelf parts to fit new or existing UVGI bulbs

The number and type of bulbs available will vary, therefore we created an easy to follow protocol to create these system utilizing almost any common BSC UV-C bulb. The design plans we developed are not limited to BSC UVGI bulbs, and we provide details to adapt them to any UVGI bulbs users have access to. The digits printed on a UVGI bulb provide the information necessary to the end-user to adapt these bulbs to create these custom UVGI lamps. For example, a bulb labeled “G30T8”. G stands for germicidal, 30 is the wattage, and T8 represents the size of the bulbs and pin geometry. For bi-pin bulbs, T5 has a ⅝ inch diameter, T8 has a 1-inch diameter, and T12 has a 1.5-inch diameter. This information is important to match to the correct fixture/bulb holder.

Similar studies analyzing the dosages required for proper UVGI decontamination and reuse of N95 masks report an optimal UV-C dose of approximately 300mJ/cm^2^ [7]. To determine UV dosage, design geometries, scalability, and N95 mask sterilization throughput of our designs we compared a model of irradiation [36] (measure of UV-C intensity per area) and compared it to measured values of UV-C irradiation from our modified fixture. UV-C intensity on a fixed plane from cylindrical source is non-linear and is a function of bulb characteristics (length,

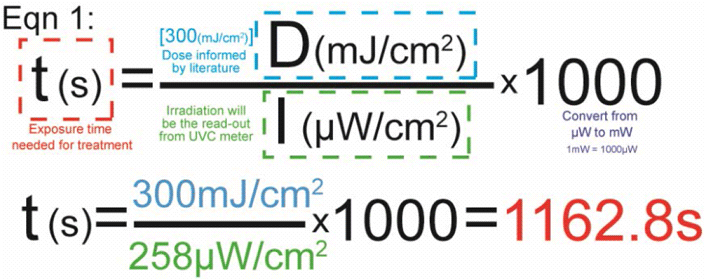

wattage, radius) and distance to the bulb (**Figure 1A**). For a single bulb, the highest irradiation will be achieved along the midline and will rapidly decay with increasing irradiation width (w) (**Figure 1B**). We used a UV meter (attenuation λ=254nm) to measure the irradiation area at discrete points using our modified fixture outfitted with a single UV-C bulb (G30T8 with 13.4W UV output) taken from an existing BSC. The bulb was 88cm long, placed 15cm above the UV-C meter sensor. We tested areas along the midline and 20cm away from the bulb’s midline in either direction. The highest irradiation was along the midline directly under the bulb and the UV intensities decay, as expected, with increasing irradiation width (w) (**Figure 1B, C)**. Importantly, these measurements demonstrate two points. Firstly, the model is a conservative prediction of the intensity distribution, and thus can be used to inform bulb configuration geometries described herein. Secondly, that the bulb intensity is not uniform along its length. Both of these details reinforce the need for user-developed designs and configurations to be validated with UV-C meter measurements to determine the minimum values of UV intensity over the exposure area to calculate exposure times needed in individual configurations.

**Figure 1:**
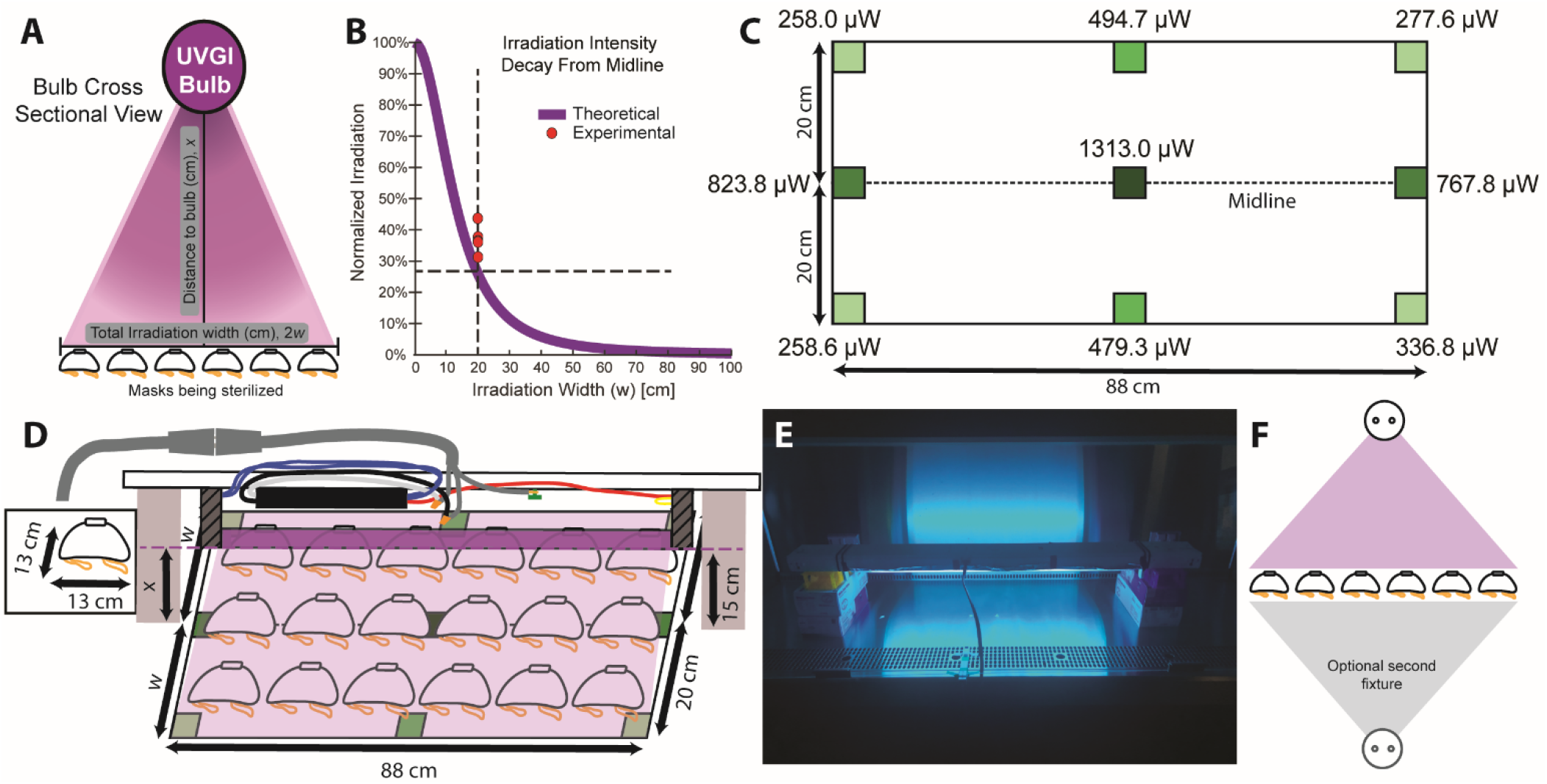
Generalized sterilization set-up options. **A**) The intensity of UV-C output is a function of bulb parameters and distance to the bulb (x), and the total irradiation width (2w). UV intensity should be validated with a UV-C meter. **B**) Theoretical UV-C irradiation decays non-linearly as distance (w) increases. **C)** Measured UV-C intensities across an irradiation field using a modified off-the-shelf light fixture using a 30W UV-C bulb, x=15cm away from the bulb. **D**) Cartoon of this particular sterilization configuration and **E**) picture of modified UVGI light. **F**) Using an optional second fixture, one can irradiate both sides of the masks simultaneously to increase mask sterilization throughput by 2x.

We measured an N95 mask (model 7048, 3M) to be 13cm wide, and therefore we could sterilize an ar ray of 6 × 3 masks (**Figure 1D, E**). Masks placed too close together can create shadows which prevents effective UV sterilization. For an array of 6 × 3 masks, using a single bulb, we use the lowest measured UV-C value over the exposure area (258.0 µW/cm^2^, from **Figure 1C**) to determine the irradiation time. To calculate the UV-C exposure time needed for N95 treatment (**Eqn 1**), a simple equation is used where treatment time, t, in seconds is calculated by dividing the desired dosage value, D, (300 mJ/cm^2^ for SARS-CoV-2 [7]) and the measured irradiance value, I, from the UV-C meter. For our measured set-up, treatment time is 1162.8 seconds or 19.4 minutes for each side of the mask.

If we reduce the distance to the bulb (x), the measured irradiation on the masks will increase non-linearly and will reduce the time needed to achieve a dose of 300mJ/cm^2^ per side of the mask. Further increases in mask sterilization throughput could be achieved by adding a second light fixture which will allow exposure to both sides of the mask simultaneously, similar the Nebraska Medicine configuration, reducing the total time for irradiation in half (**Figure 1F**).

Additional light fixtures can also be placed in parallel to increase the total irradiation area. If placed in close enough proximity, the two light fixtures will create an irradiation overlap region which will increase the irradiation intensity to reduce the time needed to sterilize (**Figure 2A**). By modeling the theoretical irradiation curves following thermal radiation view factors [36], we can determine the effects of adjusting the bulb spacing on the mask sterilization throughput capacity. For our single bulb set-up, using irradiation width w=20cm, processing 18 masks in an array, the sterilization time is 19.4 minutes per side and thus can sterilize 27.9 masks/hr (**Figure 2B**). This time is determined by the lowest intensity measured in our array where the intensity is approximately 26% that at the midline. However, if we use a modified commercial light fixture outfitted with two UV bulbs, the bulbs are closely spaced resulting in high overlap in UV light and the peak intensity of UV is notably increased (**Figure 2C**). This geometry can either produce a larger (wider) irradiation width for the same exposure time from 20cm to 29cm to increase mask throughput, or we can keep the irradiation width the same and yield a higher UV intensity. A higher UV intensity decreases the exposure time required and increases mask throughput. Although counterintuitive, **Figure 2C** demonstrates highest mask throughput (59.6 masks/hr compared to 37.2 masks/hr) by keeping the irradiation field constant (i.e. fewer masks at once) and having a higher UV intensity to decrease exposure time. These measurements and calculations are for a single modified, existing, commercial two-bulb light fixture.

**Figure 2:**
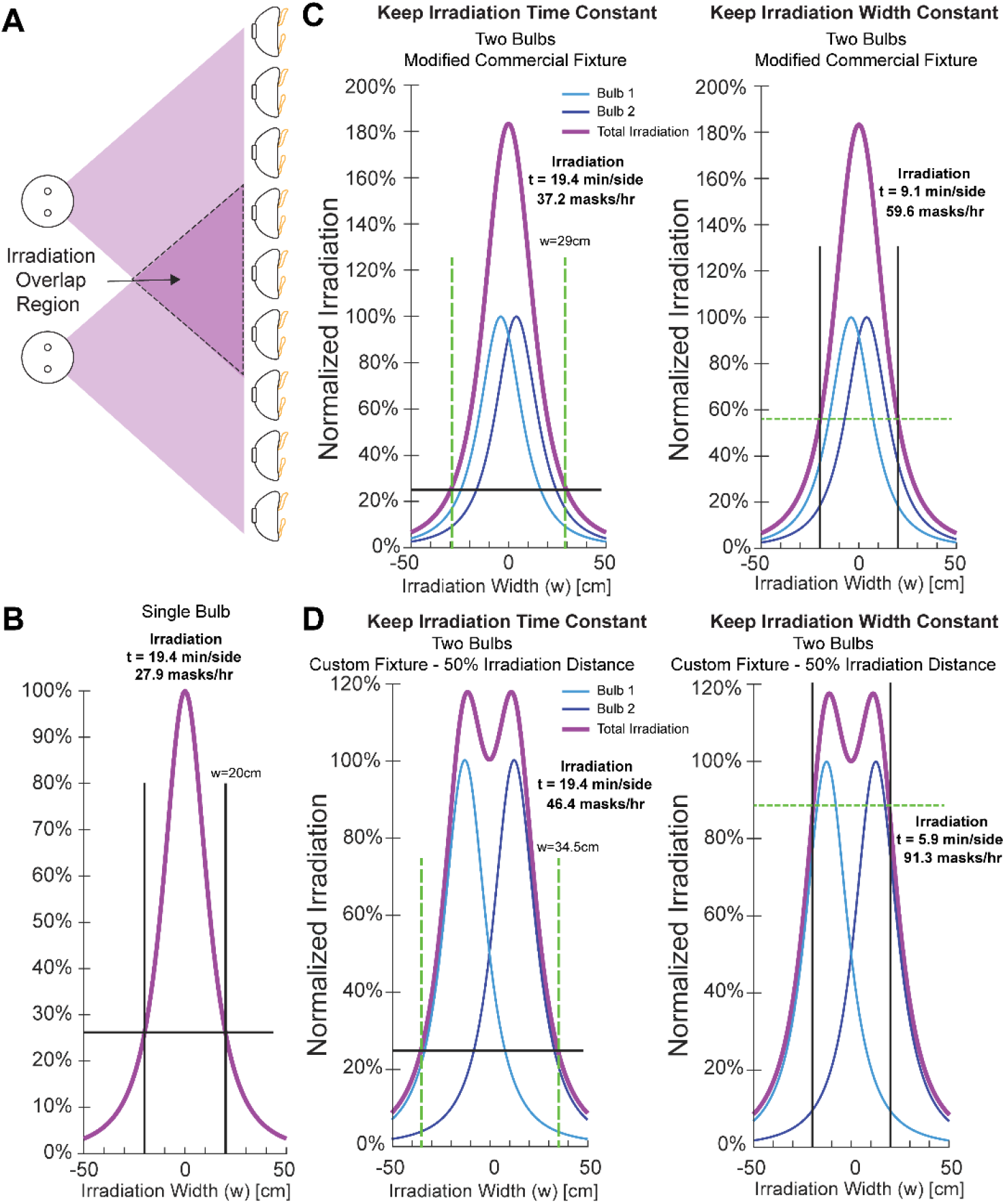
Multiple bulb arrays can improve throughput. **A**) Placing two bulbs in close proximity creates an irradiation overlap region where UV-C output is additive. **B**) Calculated irradiation curve for a single bulb Calculated UV intensity profiles of two UVGI bulbs fit into **C**) a modified off-the-shelf 2 bulb fixture or **D**) two bulbs in a custom fixture with bulbs spaced at 50% intensity irradiation width (w) for a single bulb. These calculations demonstrate the pros and cons of changing the size of the irradiation area or keeping the same area and leveraging the higher minimum intensity on mask sterilization throughput.

While having two bulbs in a commercial fixture increases throughput over a single bulb, taking advantage of off-the-shelf components to build your own custom fixture has significant advantages. Specifically, users can optimize bulb spacing to increase the uniformity of the irradiation field. If two bulbs are spaced at the 50% irradiation width intensity for a single bulb, the UV intensities are additive which creates a more constant irradiation field (**Figure 2D**). Using this configuration, the irradiation width is considerably increased from 20cm to 34.5cm for a mask exposure time of 19.4 minutes/side. This generates a mask sterilization throughput of 46.4 masks per hour. However, similar to findings from the off-the-shelf fixture, keeping the irradiation area constant to take advantage of the higher UV intensities, will increase mask throughput more substantially. This approach reduces sterilization time to 5.9 minutes per side, and yields a throughput of 91.3 masks/hr for a system with lights only on one side of the masks. This represents a greater than 3x increase in mask processing from a single UVGI bulb. If a system is created with simultaneous exposure to front and back sides of the masks, using two UVGI bulbs on each side, a user could process 182.6 masks/hr. Choosing an optimal set up will all depend on the resources at hand, such as fixtures, bulbs, and treatment environments. A significant advantage of this system is the potential for parallelization of numerous UVGI light fixtures to simply scale the N95 mask treatment throughput.

This system leverages affordable components and resources already available and distributed across the country in the variety of dif ferent research centers. Using our approach to modify commercial fixtures from the hardware store, a single bulb, two-bulb, and four-bulb UVGI light fixtures can each be constructed for less than US$25, $30, and $45, respectively. Each custom built fixture (**Figure 3A**) can be constructed for less than US$21 for a two-bulb configuration and less than US$36 for a four-bulb configuration. To implement this UVGI light source, a UV-C meter should be used to provide an accurate measurement of irradiance (µW/cm^2^) at the position of the N95 masks will be placed away from the UVGI system [4,23]. We recommend using a similar workflow and arrangement that Nebraska Medicine developed with their UVGI light towers. Both modified light fixtures or custom fixtures can be easily propped over the sterilization surface by simply resting on boxes (**Figure 1E**), or affixed to common items in a medical facility such as under a table or an IV pole, to allow customization and adaptability of the sterilization area (**Figure 3B**). Using these modified light sources adds significant flexibility in the positioning of UV-C systems to generate multiple larger-scale UVGI bulb arrays or several treatment systems in parallel.

**Figure 3:**
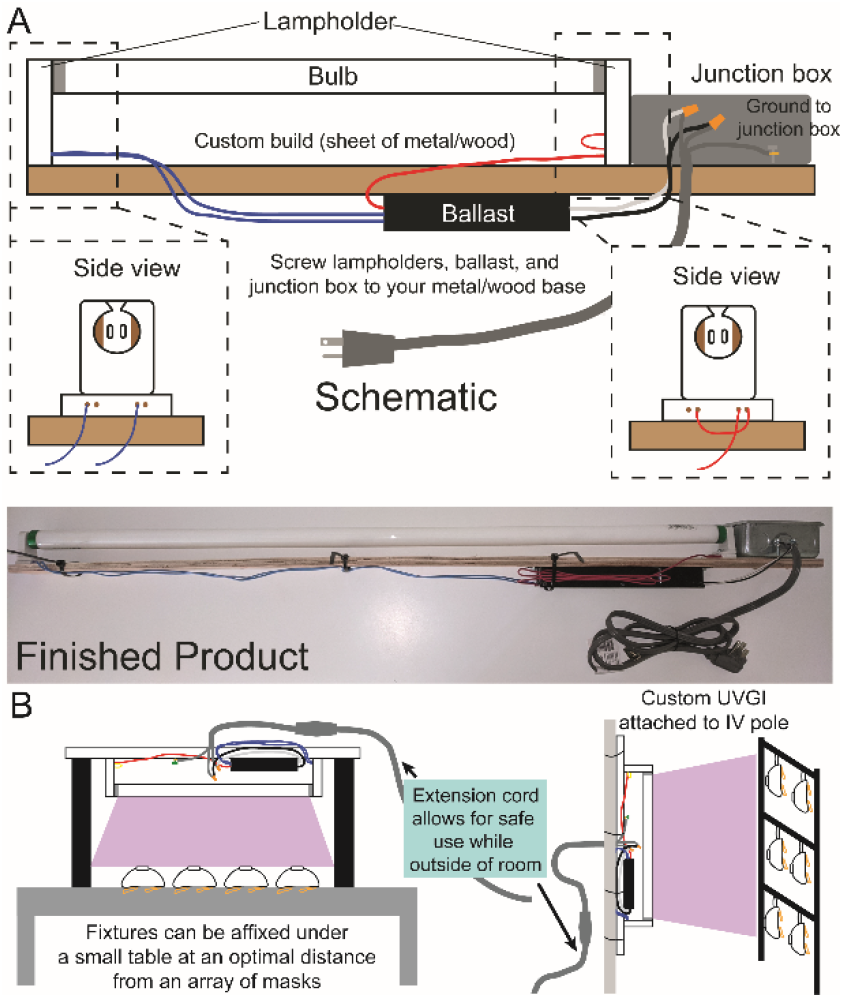
**A**) Schematic and finished product of the custom fixture that can be easily created to fit an existing UV-C bulb from a BSC. This custom fixture is one of the options depending on the end-user circumstances. The UV-C output should be measured using a UV-C meter for each individual configuration of sterilization set-up to determine the appropriate irradiation time. **B**) UVGI devices can be arrayed in a medical setting by affixing to available items such as tables or IV poles.

Our system leverages a collaboration between research institutions and hospitals, but otherwise has minimal cost and expertise associated. Research institutions can donate UVGI bulbs, UV personal protective equipment including goggles and face shields, along with the necessary UV-C meter, while hospitals can assemble the UVGI systems and implement a sterilization protocol using that established by Nebraska Medicine as an excellent starting point [7]. Our website (https://gleghornlab.com/uvgi-sterilization) provides the step-by-step downloadable plans to create the UVGI system, as well as a helpful graphics to assist end-users to determine their required sterilization times based on the measured UV-C output (λ=254nm) at a defined distance. Whereas these instructions are illustrated using materials available in the U.S., this concept has global application. The ability for front line personnel to quickly disinfect PPE on-site during any healthcare emergency could be one important measure to reduce the spread of infectious diseases, especially when time and other resources are limited.

## Data Availability

The authors confirm that the data supporting the findings of this study are available within the article and its supplementary materials.

https://gleghornlab.com/uvgi-sterilization

## Acknowledgements

While this specific project was not directly funded by specific grants, we would like to thank the funding agencies of our other projects including the National Institutes of Health, the National Science Foundation, and the Bernard Cannavan Award.

## References

1. CDC. Coronavirus Disease 2019 (COVID-19) [Internet]. Centers for Disease Control and Prevention. 2020 [cited 2020 Apr 9]. Available from: https://www.cdc.gov/coronavirus/2019-ncov/hcp/ppe-strategy/decontamination-reuse-respirators.html

2. Commissioner O of the. Coronavirus (COVID-19) Update: FDA Issues Second Emergency Use Authorization to Decontaminate N95 Respirators [Internet]. FDA. FDA; 2020 [cited 2020 Apr 18]. Available from: https://www.fda.gov/news-events/press-announcements/coronavirus-covid-19-update-fda-issues-second-emergency-use-authorization-decontaminate-n95

3. Commissioner O of the. Coronavirus (COVID-19) Update: FDA Issues Emergency Use Authorization to Decontaminate Millions of N95 Respirators [Internet]. FDA. FDA; 2020 [cited 2020 Apr 18]. Available from: https://www.fda.gov/news-events/press-announcements/coronavirus-covid-19-update-fda-issues-emergency-use-authorization-decontaminate-millions-n95

4. Padilla M. ‘It Feels Like a War Zone’: Doctors and Nurses Plead for Masks on Social Media. The New York Times [Internet]. 2020 Mar 19 [cited 2020 Apr 5]; Available from: https://www.nytimes.com/2020/03/19/us/hospitals-coronavirus-ppe-shortage.html

5. Goodnough A. Some Hospitals Are Close to Running Out of Crucial Masks for Coronavirus. The New York Times [Internet]. 2020 Mar 9 [cited 2020 Apr 5]; Available from: https://www.nytimes.com/2020/03/09/health/coronavirus-n95-face-masks.html

6. Nierenberg A. Where Are All the Masks? The New York Times [Internet]. 2020 Apr 3 [cited 2020 Apr 5]; Available from: https://www.nytimes.com/article/face-masks-coronavirus.html

7. Lowe JJ, Paladino KD, Farke JD, Boulter K, Cawcutt K, Emodi M, Gibbs S, Hankins R, Hinkle L, Micheels T, Schwedhelm S, Vasa A, Wadman M, Watson S, Rupp ME. N95 Filtering Facepiece Respirator Ultraviolet Germicidal Irradiation (UVGI) Process for Decontamination and Reuse. 2020 Mar;19.

8. Tseng C-C, Li C-S. Inactivation of viruses on surfaces by ultraviolet germicidal irradiation. J Occup Environ Hyg. 2007 Jun;4(6):400–5.

9. Mills D, Harnish DA, Lawrence C, Sandoval-Powers M, Heimbuch BK. Ultraviolet germicidal irradiation of influenza-contaminated N95 filtering facepiece respirators. Am J Infect Control. 2018;46(7):e49–55.

10. Heimbuch B, Harnish D. Research to Mitigate a Shortage of Respiratory Protection Devices During Public Health Emergencies. Applied Research Associates; 2019 Sep p. 275. Report No.: HHSF223201400158C.

11. Fast Facts on U.S. Hospitals, 2020 | AHA [Internet]. [cited 2020 Apr 4]. Available from: https://www.aha.org/statistics/fast-facts-us-hospitals

12. U.S. Cities Factsheet | Center for Sustainable Systems [Internet]. [cited 2020 Apr 4]. Available from: http://css.umich.edu/factsheets/us-cities-factsheet

13. Kenya Master Health Facility List: Find all the health facilities in Kenya [Internet]. [cited 2020 Apr 18]. Available from: http://kmhfl.health.go.ke/#/facility_filter/results

14. World Health Organization. Primary Health Care Systems (PRIMASYS): Case study from Kenya. Geneva; 2017.

15. Medical doctors (per 10 000 population) [Internet]. [cited 2020 Apr 18]. Available from: https://www.who.int/data/maternal-newborn-child-adolescent/monitor

16. Africa cannot afford to lose doctors to COVID-19 [Internet]. World Economic Forum. [cited 2020 Apr 18]. Available from: https://www.weforum.org/agenda/2020/04/africa-cannot-lose-doctors-covid-19/

17. ‘Out here, it’s just me’: In the medical desert of rural America, one doctor for 11,000 square miles - The Washington Post [Internet]. [cited 2020 Apr 4]. Available from: https://www.washingtonpost.com/national/out-here-its-just-me/2019/09/28/fa1df9b6-deef-11e9-be96-6adb81821e90_story.html

18. N95 mask shortage comes down to this key material: “The supply chain has gotten nuts” - CBS News [Internet]. [cited 2020 Apr 18]. Available from: https://www.cbsnews.com/news/n95-mask-shortage-melt-blown-filters/

19. Institute of Medicine. Reusability of Facemasks During an Influenza Pandemic: Facing the Flu. [Internet]. Washington, DE: The National Academies Press; 2006. Available from: https://doi.org/10.17226/11637

20. Carias C, Rainisch G, Shankar M, Adhikari BB, Swerdlow DL, Bower WA, Pillai SK, Meltzer MI, Koonin LM. Potential Demand for Respirators and Surgical Masks During a Hypothetical Influenza Pandemic in the United States. Clin Infect Dis. 2015 May 1;60(suppl_1):S42–51.

21. Blachere FM, Lindsley WG, McMillen CM, Beezhold DH, Fisher EM, Shaffer RE, Noti JD. Assessment of influenza virus exposure and recovery from contaminated surgical masks and N95 respirators. J Virol Methods. 2018;260:98–106.

22. Viscusi DJ, Bergman MS, Eimer BC, Shaffer RE. Evaluation of five decontamination methods for filtering facepiece respirators. Ann Occup Hyg. 2009 Nov;53(8):815–27.

23. Battelle cleared to sterilize N95 masks at max capacity, operate in other states to fight coronavirus PPE shortage | NBC4 WCMH-TV [Internet]. [cited 2020 Apr 4]. Available from: https://www.nbc4i.com/community/health/coronavirus/gov-dewine-fda-limits-battelles-ppe-mask-sterilizing-technology-to-only-10k-a-day/

24. Battelle CCDS Critical Care Decontamination System for PPE Decontamination [Internet]. Battelle. [cited 2020 Apr 4]. Available from: https://inside.battelle.org/blog-details/keeping-the-human-connection-in-drug-delivery-device-development

25. Fischer R, Morris DH, Doremalen N van, Sarchette S, Matson J, Bushmaker T, Yinda CK, Seifert S, Gamble A, Williamson B, Judson S, Wit E de, Lloyd-Smith J, Munster V. Assessment of N95 respirator decontamination and re-use for SARS-CoV-2. medRxiv. 2020 Apr 15;2020.04.11.20062018.

26. Heimbuch BK, Wallace WH, Kinney K, Lumley AE, Wu C-Y, Woo M-H, Wander JD. A pandemic influenza preparedness study: use of energetic methods to decontaminate filtering facepiece respirators contaminated with H1N1 aerosols and droplets. Am J Infect Control. 2011 Feb;39(1):e1–9.

27. Viscusi DJ, Bergman MS, Novak DA, Faulkner KA, Palmiero A, Powell J, Shaffer RE. Impact of three biological decontamination methods on filtering facepiece respirator fit, odor, comfort, and donning ease. J Occup Environ Hyg. 2011 Jul;8(7):426–36.

28. Bergman MS, Viscusi DJ, Palmiero AJ, Powell JB, Shaffer RE. Impact of Three Cycles of Decontamination Treatments on Filtering Facepiece Respirator Fit. 2011;28(1):12.

29. Bergman MS, Viscusi DJ, Heimbuch BK, Wander JD, Sambol AR, Shaffer RE. Evaluation of Multiple (3-Cycle) Decontamination Processing for Filtering Facepiece Respirators. J Eng Fibers Fabr. 2010 Dec;5(4):155892501000500.

30. Lore MB, Heimbuch BK, Brown TL, Wander JD, Hinrichs SH. Effectiveness of three decontamination treatments against influenza virus applied to filtering facepiece respirators. Ann Occup Hyg. 2012 Jan;56(1):92–101.

31. Lindsley WG, Martin SB, Thewlis RE, Sarkisian K, Nwoko JO, Mead KR, Noti JD. Effects of Ultraviolet Germicidal Irradiation (UVGI) on N95 Respirator Filtration Performance and Structural Integrity. J Occup Environ Hyg. 2015;12(8):509–17.

32. Anderson JG, Rowan NJ, MacGregor SJ, Fouracre RA, Farish O. Inactivation of food-borne enteropathogenic bacteria and spoilage fungi using pulsed-light. IEEE Trans Plasma Sci. 2000 Feb;28(1):83–8.

33. van Doremalen N, Bushmaker T, Morris DH, Holbrook MG, Gamble A, Williamson BN, Tamin A, Harcourt JL, Thornburg NJ, Gerber SI, Lloyd-Smith JO, de Wit E, Munster VJ. Aerosol and Surface Stability of SARS-CoV-2 as Compared with SARS-CoV-1. N Engl J Med. 2020 Mar 17;0(0):null.

34. Mamahlodi MT. Potential benefits and harms of the use of UV radiation in transmission of tuberculosis in South African health facilities. J Public Health Afr [Internet]. 2019 Jun 12 [cited 2020 Apr 18];10(1). Available from: https://www.ncbi.nlm.nih.gov/pmc/articles/PMC6589622/

35. Card KJ, Crozier D, Dhawan A, Dinh M, Dolson E, Farrokhian N, Gopalakrishnan V, Ho E, King ES, Krishnan N, Kuzmin G, Maltas J, Pelesko J, Scarborough JA, Scott JG, Sedor G, Weaver DT. UV Sterilization of Personal Protective Equipment with Idle Laboratory Biosafety Cabinets During the Covid-19 Pandemic. medRxiv. 2020 Mar 27;2020.03.25.20043489.

36. Kowalski W. Ultraviolet Germicidal Irradiation Handbook. Springer International Publishing; 2009. 493 p.

